# Premature infants display discriminable behavioural, physiological and brain responses to noxious and non-noxious stimuli

**DOI:** 10.1101/2021.08.18.21262106

**Authors:** Marianne van der Vaart, Caroline Hartley, Luke Baxter, Gabriela Schmidt Mellado, Foteini Andritsou, Maria M. Cobo, Ria Evans Fry, Eleri Adams, Sean Fitzgibbon, Rebeccah Slater

## Abstract

Pain assessment in preterm infants is challenging, as behavioural, autonomic and neurophysiological measures of pain are reported to be less sensitive and specific than in term infants. Understanding the pattern of preterm infants’ noxious-evoked responses is vital to improve pain assessment in this group. This study investigated the discriminability and development of multi-modal noxious-evoked responses in infants aged 28-40 weeks postmenstrual age. A classifier was trained to discriminate responses to a noxious heel lance from a non-noxious control in 47 infants, using measures of facial expression, brain activity, heart rate and limb withdrawal, and tested in two independent cohorts with a total of 98 infants. The model discriminates responses to the noxious from the non-noxious procedure from 28 weeks onwards with an overall accuracy of 0.77-0.83 and an accuracy of 0.78-0.79 in the 28-31 week group. Noxious-evoked responses have distinct developmental patterns. Heart rate responses increase in magnitude with age, while noxious-evoked brain activity undergoes three distinct developmental stages, including a previously unreported transitory stage consisting of a negative event-related potential between 30-33 weeks postmenstrual age. These findings demonstrate that while noxious-evoked responses change across early development, infant responses to noxious and non-noxious stimuli are discriminable from 28 weeks onwards.

## Introduction

The measurement and treatment of pain in the neonatal intensive care unit (NICU) is vital to prevent short- and long-term consequences of pain in neonates (McPherson et al. 2020). However, the nervous system undergoes rapid changes in structure and function across the preterm period and observed responses to noxious stimuli evolve accordingly (Fitzgerald 2005; Hatfield 2014). When the term-aged neonate undergoes a painful medical procedure, such as a blood test, cannulation or injection, distinct noxious-evoked activity can be observed across multiple levels of the nervous system. For example, a noxious procedure can evoke autonomic responses, including increased heart rate (Waxman et al. 2016) and skin conductance (Eriksson et al. 2008), spinally mediated limb withdrawal activity (Cornelissen et al. 2013; Hartley et al. 2015), and behavioural facial grimacing responses (Grunau et al. 1990; DiLorenzo et al. 2018). In addition, patterns of noxious-evoked brain activity that are distinct from that evoked by non-noxious sensory stimulation have been well-characterised (Slater et al. 2010; Hartley et al. 2017).

In contrast, in the premature neonate the observed responses following painful medical procedures are reported to be less able to discriminate noxious from non-noxious events. For example, a similar percentage of very preterm infants display facial grimacing in response to either heel lancing or non-noxious touch (Green et al. 2019); similar magnitude limb withdrawal responses can occur in response to noxious and non-noxious stimuli (Cornelissen et al. 2013); and some studies report that heart rate responses are smaller or even absent in extremely preterm infants compared with term infants (Craig et al. 1993; Porter et al. 1999), although others report no relationship between heart rate response to noxious stimuli and age in preterm infants (Walden et al. 2001; Bartocci et al. 2006). Similarly, the distinct noxious-evoked brain activity patterns observed in term-aged neonates are less frequently observed in younger infants, and in very premature infants generalised delta brush activity is observed in response to both noxious and non-noxious sensory events (Fabrizi et al. 2011). Consequently, the sensitivity of clinical pain scales in the youngest infants has been questioned, leading to limitations in using these outcomes to measure pain or to assess the efficacy of analgesics in this population (Slater et al. 2020).

Nevertheless, observations that noxious and non-noxious stimulation evoke similar response patterns in premature neonates should not be interpreted to mean that the neonatal nervous system does not discriminate noxious from non-noxious events. Animal studies show that subtypes of A-fibres which are differentially activated by noxious and tactile inputs are functional at birth in rodents (Brewer and Baccei 2020) a developmental stage which corresponds to late preterm infant development. This suggests that the preterm infant nervous system may also be able to generate discriminable responses during this time window.

Therefore, depending on the age of the infant, it may be that we cannot discriminate between noxious- and tactile-evoked responses because the evoked patterns of brain activity are truly not discriminable between these events (true negative) or, alternatively, that we cannot observe discriminable responses across these modalities due to limitations in our measurement techniques, despite the responses being able to discriminate (false negative). Given that each technique provides relatively crude measures of central nervous system (CNS) activity due to the necessity that they are non-invasive and the essential requirement that they are clinically acceptable for use in the neonate, there is an abundance of methodological limitations that could lead to a failure to reject the null hypothesis. For example, evoked electroencephalography (EEG) signals are susceptible to artefact arising from infant movement (Hoehl and Wahl 2012) and can have a low signal-to-noise ratio in single trials (Boudewyn et al. 2018). Utilising multiple recording modalities and measurement types in premature infants may be one approach whereby we can improve our power to discriminate responses to noxious and non-noxious stimuli, as has previously been shown in late preterm and term infants (Zamzmi et al. 2017; van der Vaart et al. 2019).

The aim of this study was to determine whether infants between 28-40 weeks postmenstrual age (PMA) display discriminable physiological, behavioural, reflexive, and cerebral responses to noxious and non-noxious events, and if so, to investigate how these responses change throughout early human development.

## Materials and methods

### Study design

The study consisted of two parts that related to the two main aims:

1. Part 1: Discrimination: investigation of the discriminability between responses to noxious and non-noxious stimulation.
2. Part 2: Development: investigation of the age-related development of noxious-evoked responses across 28-40 weeks PMA.

A total of 145 infants were included in this study in three datasets (Table 1). The local ‘Oxford’ dataset (n = 71) consists of the Oxford Training Dataset (67% of Oxford participants, n = 47) and an independent Oxford Held-out Test Dataset (33% of Oxford participants, n = 24). The ‘UCL’ dataset (n = 74) is an external dataset compiled by Jones and colleagues at University College London (UCL) (Jones et al. 2018b, 2018a).

**Table 1.** Demographic information of participating infants, split by dataset.

|  | **Oxford Training** | **Oxford Held-out Test** | **UCL** |
| --- | --- | --- | --- |
|  | **(n = 47)** | **(n = 24)** | **(n = 74)** |
| **PMA at test occasion** | 35.3 (32.2-37.6) | 34.8 (32.3-37.2) | 35.6 (33.0-37.1) |
| **(weeks)** |  |  |  |
| **Gestational age at** | 34.3 (29.2-37.2) | 34 (29.5-36.7) | 34.3 (31.0-36.3) |
| **birth (weeks)** |  |  |  |
| **PNA at test occasion** | 0.7 (0.3-2) | 0.7 (0.2-1.6) | 0.7 (0.6-1.6) |
| **(weeks)** |  |  |  |
| **Sex** |  |  |  |
| **Females** | 21 (45) | 11 (46) | 38 (51) |
| **Males** | 26 (55) | 13 (54) | 36 (49) |
| **Birth weight (g)** | 2110 (1183-3034) | 2002 (1500-3153) | 2070 (1490, 2480) |
| ***Mode of delivery*** |  |  |  |
| **Normal vaginal delivery** | 16 (34) | 10 (42) | 19 (26) |
| **Vaginal breech** | 3 (6) | 0 (0) | 3 (4) |
| **Vaginal assisted (ventouse / forceps / kiwi)** | 6 (13) | 4 (17) | 12<sup>1</sup> (16) |
| **Elective C-section** | 4 (9) | 3 (13) | 15 (20) |
| **Emergency C-section / C-section in labour** | 18 (38) | 7 (29) | 25 (34) |
| **Apgar at 1 minute** | 8 (5.3-9) | 7 (4-9) | 8 (6-9)<sup>2</sup> |
| **Apgar at 5 minutes** | 10 (8-10) | 9.5 (8-10) | 9 (9-10)<sup>2</sup> |
| **Estimated number of previous skin-breaking blood tests** | 9 (3-16)<sup>3</sup> | 5.5 (2.5-10) | 13 (7-20) |

Values are median (lower quartile, upper quartile) or number (%). Abbreviations: PMA = postmenstrual age; PNA = postnatal age; UCL = University College London.
1. Includes one assisted vaginal breech birth. 2. Apgar scores missing for 2 infants. 3. Estimated number of previous skin-breaking blood tests missing for 2 infants.

For Part 1: Discrimination, the Oxford Training Dataset was used for feature extraction, model training and assessment of classification performance using cross-validation, while the Oxford Held-out Test Dataset and the UCL Dataset were kept separately. The model established in the Oxford Training Dataset was then tested in both the independent Oxford Held-out Test Dataset and the independent UCL Dataset. The Oxford Held-out Test Dataset was used to test the generalisability of the classification accuracy in an independent dataset collected at the same site, while the UCL dataset was used to assess generalisability across different sites. For Part 2: Development, the Oxford Training data, the Oxford Held-out Test data and UCL data were combined to maximise sample size and best represent the developmental trajectories of individual features.

#### Oxford dataset

##### Participants and research governance

Infants were selected from a database of all data previously recorded by our research group between 2012 and 2021 at the John Radcliffe Hospital, Oxford University Hospitals NHS Foundation Trust, Oxford, UK. Ethical approval was obtained from the National Research Ethics Service (references: 12/SC/0447 and 19/LO/1085) and informed written parental consent was obtained before each study. Studies conformed to the standards of the Declaration of Helsinki and Good Clinical Practice guidelines.

Infants were included in the current analyses if they were aged between 28 and 40 weeks PMA at the time of the study and if at least 1.5 seconds of time-locked artefact-free EEG was recorded at the Cz electrode during a heel lance and a control heel lance. Infants were excluded from this study if they had a postnatal age (PNA) of 8 weeks or more, intraventricular haemorrhage (IVH) grade 3 or 4, hypoxic-ischaemic encephalopathy, if their data were used in the development of the previously described template of noxious-evoked brain activity (Hartley et al. 2017) or if they were in the intervention group of a study that specifically investigated the effect of a pain-relieving intervention (e.g., gentle touch or kangaroo care). The database contained fewer preterm infants than (near-)term infants. To prevent data from (near-)term age infants dominating the dataset, a maximum of 10 infants per postmenstrual week was included by selecting study numbers from a list before analysing the data. Some infants in the database participated in multiple test occasions; only one test occasion per infant was included in this study. Parts of this dataset were previously used for other analyses (Hartley et al. 2016; Gursul et al. 2018; Green et al. 2019; van der Vaart et al. 2019; Cobo et al. 2021; Schmidt Mellado et al. 2021).

The selected infants were randomly split into the Oxford Training Dataset (n = 47) and the Oxford Held-out Test Dataset (n = 20), stratified in weeks PMA at study, using the function cvpartition in MATLAB (Mathworks, version 2019b). During feature extraction and model development in the Oxford Training Dataset, four infants fitting the inclusion criteria were recruited and were included in the Held-out Test set bringing the total infants in the Oxford Held-out Test Dataset to 24 infants. Demographic information can be found in Table 1.

### Experimental procedures

All infants were studied during a noxious stimulus – a clinically-required heel lance performed for blood sampling, and a non-noxious stimulus – a control heel lance where the lancet was rotated by 90° and held against the infant’s foot so that when released the blade did not pierce the infant’s skin. Infants received comfort techniques such as swaddling or use of a dummy or were held by a parent during the heel lance and control heel lance.

### Recording techniques

EEG was recorded using a SynAmps RT 64-channel headbox and amplifiers (Compumedics Neuroscan) and CURRYscan7 neuroimaging suite (Compumedics Neuroscan). Activity was recorded at Cz and 3 to 20 other electrodes with the reference electrode at Fz and a ground electrode at FPz/forehead according to the international 10-20 system. In two infants, data were recorded with a reference electrode at FPz and re-referenced to Fz during analysis. Sampling frequency was 2000 Hz (69 infants) or 1000 Hz (two infants, upsampled to 2000 Hz during analysis). The scalp was gently cleaned with preparation gel (Nuprep gel, D.O. Weaver and Co.) before the placement of disposable Ag/AgCl cup electrodes (Ambu Neuroline) with conductive paste (Elefix EEG paste, 8 Nihon Kohden). Electrocardiogram (ECG) was recorded with an electrode above the clavicle which was referenced to the EEG reference electrode. Electromyogram (EMG) was recorded using surface bipolar electrodes (Ambu Neuroline) on the biceps femoris of both legs. The stimuli were time-locked to the electrophysiological (EEG, EMG, and ECG) recordings with an accelerometer and an automated detection interface (Worley et al. 2012). In 2 infants the stimuli were time-locked using a microphone that recorded the click produced by the lancet and was recorded along with the electrophysiological recordings.

Infants’ facial expressions were recorded using a video camera for 15 seconds before until 30 seconds after the heel lance and control heel lance (see Facial Expressions below). The timing of the stimulus was marked with a LED-light that was activated by one of the investigators at the point of stimulation.

#### UCL dataset

The dataset from Jones and colleagues, which is available on request through the UK Data Service (Jones et al. 2018b, 2018a), contains EEG recordings and facial expression scores for 112 infants during a heel lance, control heel lance and auditory stimulus. These data were collected by an independent research group at a different site than our Oxford dataset, but using similar acquisition methods and stimuli. The UCL study was approved by the NHS Health Research Authority (London – Surrey Borders) and conformed to the declaration of Helsinki. Written parental consent was obtained before each test occasion. Further details on research governance and recording methods can be found in the associated publication (Jones et al. 2018b). A total of 74 infants in the dataset met our inclusion criteria (they had both a control heel lance and a heel lance, were aged between 28 and 40 weeks PMA, and less than 8-weeks PNA at study) and were included in our analysis. Of these infants, 9 had IVH of unknown severity. Comfort methods such as swaddling and kangaroo care were used for each test occasion. Demographic information can be found in Table 1 and age distributions are plotted in Supplementary Fig. S1.

### Analysis

#### Features of interest

Relevant literature was reviewed to identify modalities that could potentially contribute to the discrimination of noxious from non-noxious stimuli in a wide age range. Facial expressions, noxious-evoked event-related potential (ERP) magnitude and EEG spectral changes were included in the analysis because previous studies report that these modalities are more responsive to noxious stimulation than to non-noxious stimulation in term infants (Grunau et al. 1990; Slater et al. 2010; Fabrizi et al. 2016; Hartley et al. 2017). Heart rate was included as it has discriminable value in (near-) term infants (Bartocci et al. 2006; van der Vaart et al. 2019) and potentially in preterm infants (Johnston et al. 1995; Holsti et al. 2005). Limb withdrawal is reported to be present in both term and preterm infants (Hartley et al. 2016), with discriminative ability increasing with infant PMA (Cornelissen et al. 2013). As preterm infants are less likely to display the well characterised noxious-evoked ERP (Fabrizi et al. 2011), a data-driven approach using principal component analysis (PCA) (see below) was used to identify new potentially discriminable features in timelocked EEG recordings in preterm infants. Finally, PMA at test occasion was included in the model as it may modulate noxious-evoked responses. The modalities and features derived from them are described in Table 2.

**Table 2.**
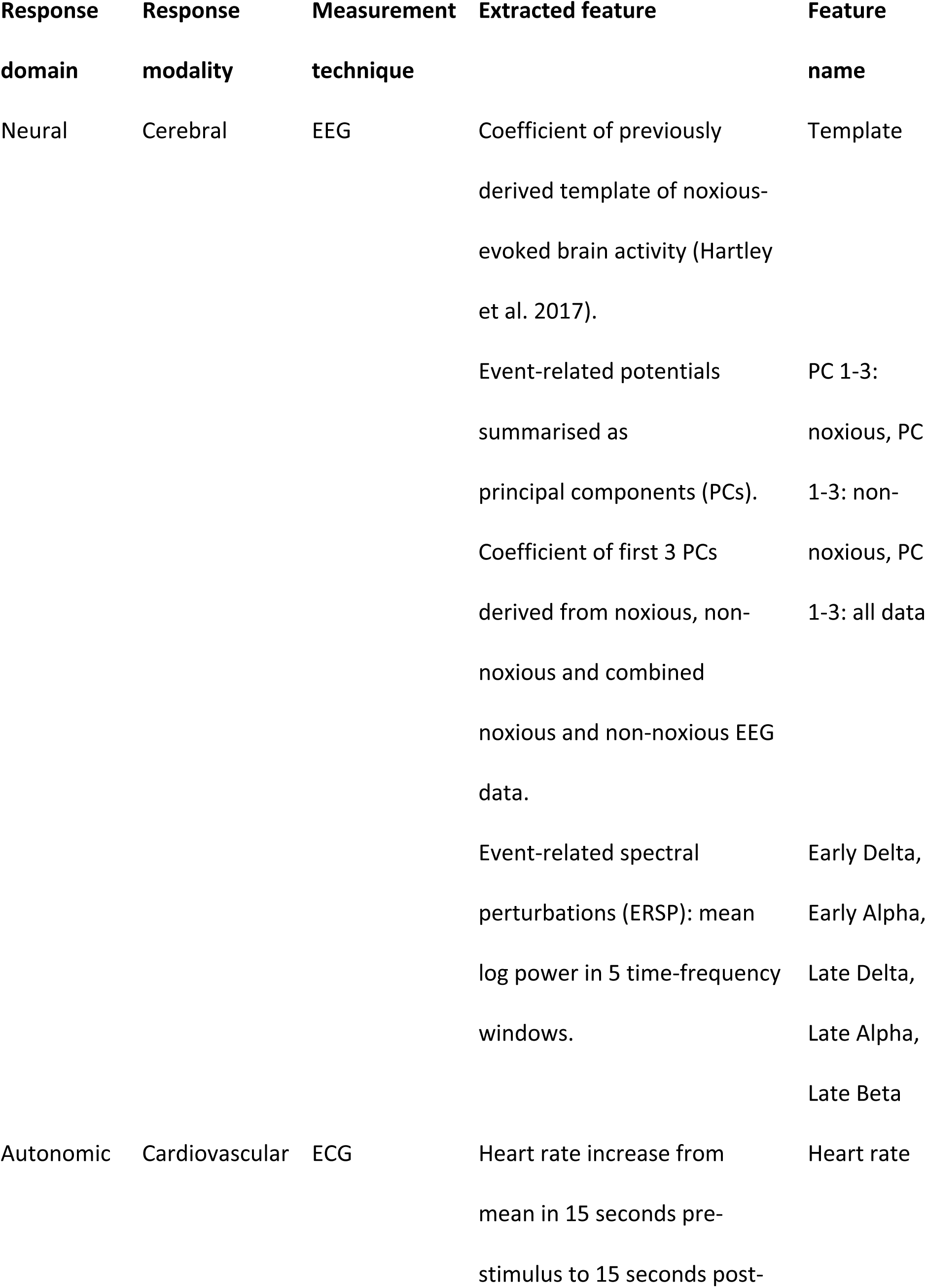
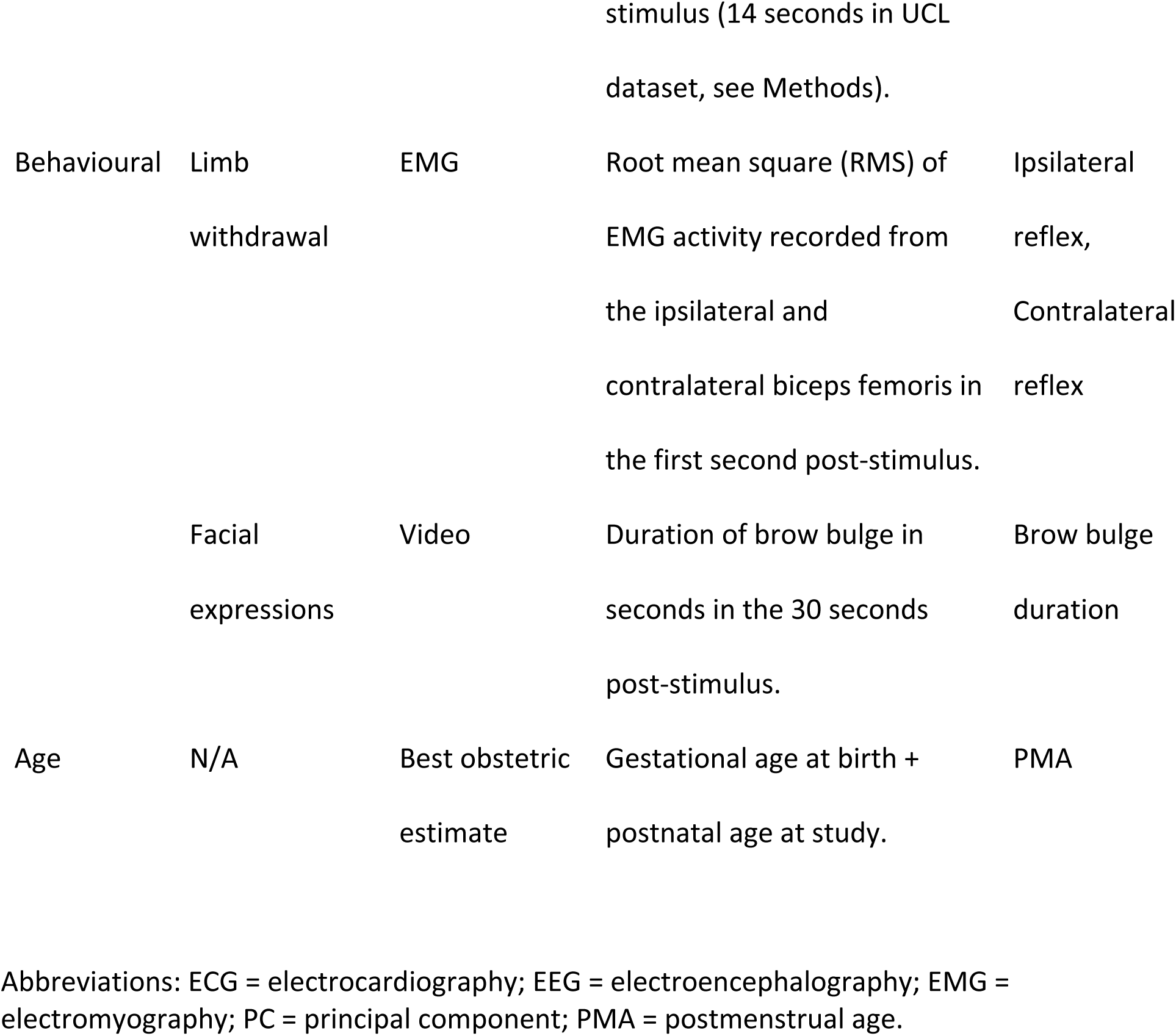
Response modalities and features used in this study, organised by response domain.

#### EEG pre-processing

EEG data were processed using Brainstorm (Tadel et al. 2011) and EEGlab (Delorme and Makeig 2004) functions in the MATLAB programming environment (Mathworks, version 2019b). Data were filtered with a low-pass filter of 30 Hz, a high pass filter of 1 Hz and a notch filter at 50 Hz using a Hamming windowed sinc FIR filter implemented in EEGlab. Data were epoched around the control heel lance and heel lance stimuli with 0.5 seconds before and 1 second after the stimulus for PCA and for projecting the previously developed template of noxious-evoked brain activity, and 2 seconds before the stimulus and 4 seconds after the stimulus for the event-related spectral perturbations (ERSP) analysis. Epochs were visually inspected and rejected if artefact was present (see Table 4).

**Table 3.** Time-frequency windows of interest.

| **Name** | **Time post-stimulus** | **Frequency** |
| --- | --- | --- |
| Early delta | 250 – 750 ms | 2 – 4 Hz |
| Early alpha | 250 – 750 ms | 7 – 15 Hz |
| Late delta | 1000 – 2000 ms | 1 – 2 Hz |
| Late alpha | 1000 – 2000 ms | 7 – 15 Hz |
| Late Beta | 1500 – 2300 ms | 28 – 30 Hz |

**Table 4.** Overview of missing data for each feature, split by dataset.

|  | Number of missing entries |  |  |
| --- | --- | --- | --- |
|  | Oxford - Training | Oxford – Held-out<br>Test | UCL |
| EEG template of<br>noxious-evoked<br>brain activity and<br>PCA (< 1 second) | 0/47 | 0/24 | 0/74 |
| EEG ERSP analysis (4<br>seconds post-stimulus) | 3/47 heel lances<br>excluded due to<br>movement artefact<br>occurring after 1<br>second. | 1/24 heel lances<br>excluded due to<br>movement artefact<br>occurring after 1<br>second. | N/A: only 2 seconds<br>of post-stimulus EEG<br>data available. |
| Heart rate | 4/47 infants excluded<br>in both conditions<br>due to ECG recording<br>artefact. | 1/24 infants excluded<br>in both conditions<br>due to presence of<br>ectopic beats. | 21/74 control heel<br>lances unavailable (3<br>due to artefact, 18<br>due to less than 14<br>seconds of ECG) and<br>9/74 heel lances<br>unavailable (7 due to<br>artefact, 1 with ECG<br>less than 14 seconds and 1 because ECG was not available). |
| **Ipsilateral reflex** | 2/47 infants excluded due to movement and recording artefact. | 0/24 | N/A: no EMG available. |
| **Contralateral reflex** | 1/47 infants excluded in both conditions due to movement and recording artefact, in 2/47 infants contralateral EMG was not recorded. | 0/24 | N/A: no EMG available. |
| **Brow bulge duration** | 3/47 infants excluded in both conditions due to failed video recording. | 2/47 infants excluded in both conditions due to failed video recording. | 4/74 heel lances and 4/74 control heel lances unavailable (data from 5 infants). |

Abbreviations: ECG = electrocardiography; EEG = electroencephalography; EMG = electromyography; PC = principal component; PCA = principal component analysis; PMA = postmenstrual age.

#### Template of noxious-evoked brain activity

A template of noxious-evoked brain activity suitable for infants aged 34-42 weeks PMA was previously developed and validated in an independent cohort of infants (Hartley et al. 2017). To calculate the weights of this template of noxious-evoked brain activity in the current datasets, EEG epochs were first baseline-corrected to the pre-stimulus mean and then the template of noxious-evoked brain activity was projected to each infant’s data as described in the original publication (Hartley et al. 2017).

#### Extraction of main ERP waveforms using PCA

In many studies, the term ERP refers to the event-related response most typically derived by averaging many trials within and across participants time-locked to a stimulus. Here we investigated responses to a clinically-required noxious stimulus (and a non-noxious control) which is necessarily a single trial and therefore required alternate methods for derivation. We use the term ERP to describe both the time-locked single-trial responses and the time-locked average responses across infants.

PCA was used to quantify the magnitude of ERPs evoked by the noxious and non-noxious stimuli. PCA permits the investigation of the entire waveform concurrently, instead of focussing on peak amplitudes. In our analysis, epochs were considered as variables and timepoints as observations. We aimed to extract the PCs that explained most of the variance in the noxious and non-noxious responses at the Cz electrode in the Oxford Training Dataset. Therefore, we used temporal PCA to extract the main waveforms from the responses to the noxious stimulus only, responses to the non-noxious stimulus only and the responses to both the noxious and non-noxious stimulus together. The procedure below was followed for each set of responses.

First, individual epochs were baseline corrected to the pre-stimulus mean. Then, each infants’ data were aligned to an age-weighted average by using Woody filtering to allow for age-related inter-individual differences in latency to evoked responses. To do so, an age-weighted average signal was calculated for each infant by using average weighting of that infant’s and the other infants’ epochs. A gaussian window with a length of 8 weeks (56 days) was used to assign weights between 0 and 1 to the other traces based on PMA at test occasion in days. This meant that weights were maximal for traces belonging to infants with the same PMA and decayed to 0.5 for traces belonging to infants who were approximately 2 weeks older or younger and to 0 for infants who were more than 4 weeks older or younger (see Supplementary Fig. S2). The infant’s signal was then aligned to this average using Woody filtering (with a maximum jitter of ±50 ms) in the first second after the stimulus. PCA was performed on the set of Woody filtered signals in the first 1000 ms post-stimulus to identify the waveforms explaining most of the variance in the data. For each set of responses (noxious, non-noxious and combined noxious and non-noxious), the first PCs that together explained 80% of the variance were considered for further analysis. This yielded three PCs per response set and nine PCs in total.

The nine PCs that were identified in the Oxford Training Dataset were projected onto all data in the Oxford dataset (Training and Held-out Test Datasets) and UCL Dataset. To do so, the PCs were decomposed using singular value decomposition and the resulting unitary matrices were multiplied with each infant’s data (Hartley et al. 2017). Each trial was first Woody filtered with a maximum jitter of ±50 ms so that the data achieved best alignment with each PC. The corresponding weights were the features that were brought forward to the classification model.

#### ERSP analysis

ERSP analysis was performed using EEGlab (Makeig 1993; Grandchamp and Delorme 2011). Data were decomposed into 200×59 time-frequency pixels using Morlet wavelets. The number of cycles was set to linearly increase from 3 cycles at 1 Hz to 45 cycles at 30 Hz, which corresponds to 50% of the number of cycles in the equivalent fast Fourier transform window (Makeig 1993). Individual trials were baseline corrected by dividing the post-stimulus power at each frequency by the average power in the same frequency band in the 500 ms baseline period directly before the stimulus (Makeig 1993; Grandchamp and Delorme 2011). Data were then log-transformed at the single-trial level.

In the average ERSP plot in the Oxford Training Dataset, we visually identified five time-frequency windows that were responsive to noxious stimulation (prominent positive or negative deviations from zero), analogous to identifying peaks in temporal EEG analysis (Fig. M1, Table 3). The mean log power in the five time-frequency windows identified in the Oxford Training Dataset were used as features in the classification model and were calculated for each individual epoch in the Oxford Training Dataset and Oxford Held-out Test Dataset. The UCL Dataset only contained 2 seconds of post-stimulus EEG recordings which was too short to calculate the time-frequency decomposition as described above.

**Figure M1.**
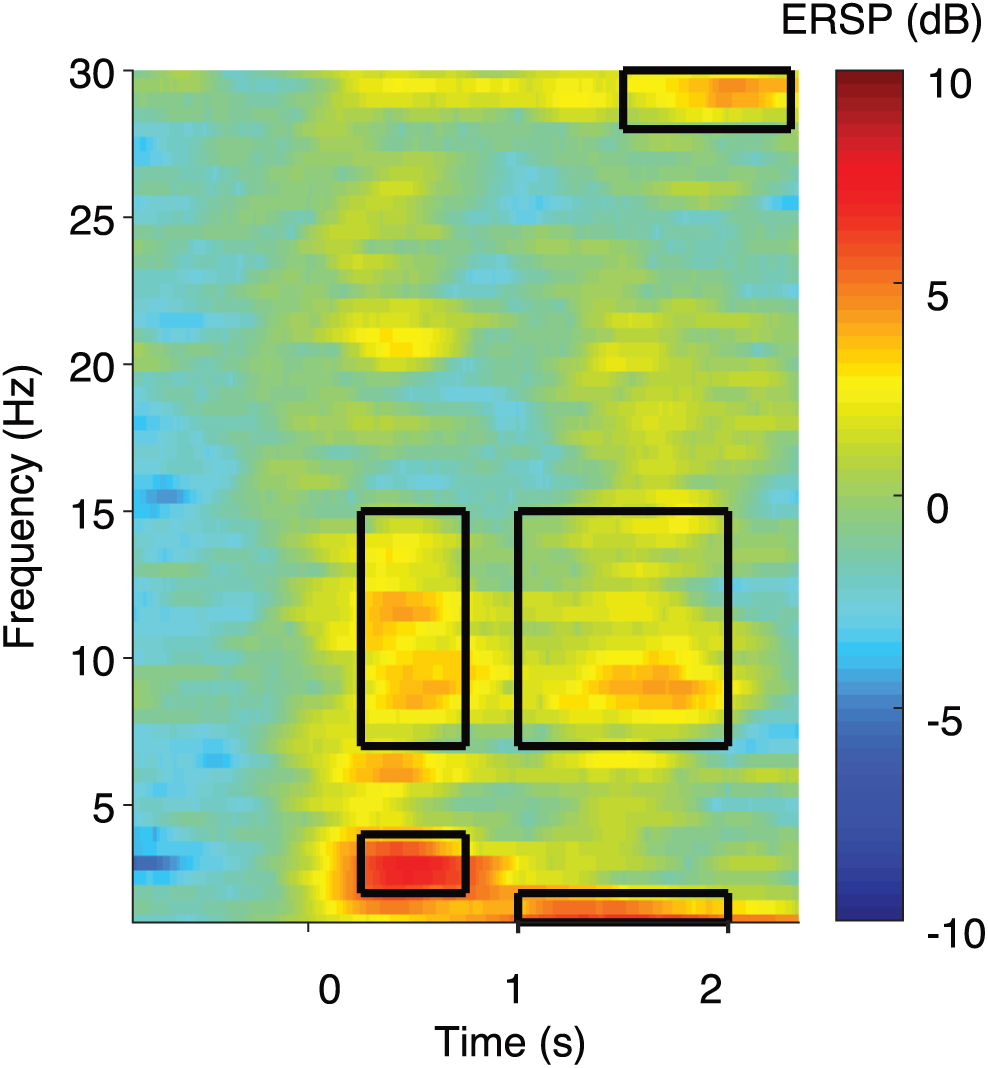
Mean ERSP plot in response to the noxious stimulus in the Oxford Training Dataset. Boxes indicate the time-frequency windows of interest identified using this plot. The noxious stimulus (clinically required heel lance) was performed at time = 0. Abbreviations: ERSP = event-related spectral perturbations.

#### ECG pre-processing and feature extraction

ECG data were filtered between 12 and 40 Hz using a Hamming windowed sinc FIR filter implemented in EEGlab (Delorme and Makeig 2004) and divided into epochs with 30 seconds pre- and post-stimulus for the Oxford data and 14 seconds pre- and post-stimulus for the UCL dataset (as 14 seconds of data was available for most infants in this dataset). The MATLAB function findpeaks.m was used to identify R peaks; traces were visually examined and extra or missed peaks were manually corrected. Traces with severe artefact were rejected (see Table 4). Heart rate was calculated at every second by taking the inverse of the average RR interval in 3-second sliding windows. Mean heart rate in the 15 seconds before the stimulus was calculated and subtracted from the maximum heart rate in 15 seconds post-stimulus, as this was shown to be a feature that could discriminate noxious from non-noxious stimulus responses in a previous model developed in infants from 34 weeks PMA (van der Vaart et al. 2019), and this was used as the heart rate feature in the classification model. For the UCL Dataset, the mean in the 14 seconds pre-stimulus was subtracted from the maximum in the 14 seconds post-stimulus.

#### EMG pre-processing and feature extraction

EMG data were rectified, filtered between 10 and 500 Hz with a notch filter at 50 Hz and harmonics using a Hamming windowed sinc FIR filter implemented in EEGlab (Delorme and Makeig 2004), and epoched with 5 seconds before the stimulus and 15 seconds after the stimulus. Data were visually inspected to identify artefacts and removed if necessary (see Table 4).

The root mean square (RMS) of the signal was calculated in 250 ms bins and the post-stimulus RMS data were divided by the mean RMS in the one second pre-stimulus (to correct for inter-individual differences in baseline EMG signal). The mean baseline-corrected RMS in the first 1000 milliseconds post-stimulus was used as a feature in the classification model. This time window was chosen as EMG RMS activity in the first 1000 ms is differentially modulated by noxious and non-noxious stimuli in term infants (Hartley et al. 2015), is more comparable between term and preterm infants than later time windows (Cornelissen et al. 2013) and is less likely to be contaminated by other behavioural responses, which tend to occur after 1 second (Slater et al. 2009).

#### Facial expressions

The duration of brow bulge in seconds in the 30 seconds after the control heel lance and heel lance was scored according to the feature within the Premature Infant Pain Profile – Revised (PIPP-R) score (Stevens et al. 2014) by researchers trained in this scoring system. For the Oxford Dataset, researchers were blinded to the stimulus type. For the UCL Dataset, scoring was performed by the original research team in UCL and we obtained the facial expression duration as part of the dataset that was shared (see (Jones et al. 2018b) for details). Brow bulge was chosen as a feature for our model as it has been previously shown to discriminate noxious from non-noxious stimuli (van der Vaart et al. 2019).

#### Missing data

Not all variables were available for all infants due to technical difficulties during recordings or artefacts. The UCL Dataset did not contain EMG data. Table 4 summarises missing data.

#### Part 1: Discrimination - Classification model

To investigate the ability to discriminate noxious from non-noxious responses across the age range from 28-40 weeks PMA using multimodal responses and PMA, we used a bagged decision trees classification model (Breiman et al. 1984; Breiman 2001). The bagged decision tree model was chosen because it provides robust predictions and can handle missing entries in the data (García-Laencina et al. 2010). The model was implemented in MATLAB using the function TreeBagger, with 500 weak learners and a minimum leaf size of 3, to classify responses as either noxious (heel lance) or non-noxious (control heel lance). The model used 20 features (see Table 2): nine EEG PC features obtained through PCA, the weight of the previously described template of noxious-evoked brain activity (Hartley et al. 2017), mean log power increase in the 5 time-frequency windows identified in the ERSP analysis, increase in heart rate, ipsilateral and contralateral limb withdrawal quantified by EMG RMS, the duration of brow bulge and PMA at study. Surrogate splits were specified to improve predictions in data with missing variables and the interaction-curvature test was used to allow for interactions between pairs of variables and to obtain unbiased estimates of predictor importance. All the predictors could be selected at each split (in contrast to random forest) to get unbiased estimates of predictor importance.

The model was trained in the Oxford Training Dataset using leave-one-infant out cross-validation (i.e., responses to both the noxious and non-noxious stimulus for a single infant were left out) to get an estimate of performance in the training set. The model was then retrained on all the data in the Oxford Training Dataset and this final model was tested on the two independent datasets, the Oxford Held-out Test Dataset and the UCL Dataset, to assess generalisability. The UCL Dataset only contained temporal EEG features, heart rate increase and brow bulge duration and all other features were considered missing data. Classification accuracy with 95% binomial proportion confidence interval was used to assess model performance. Given the equal number of observations for the noxious and non-noxious categories, this was a balanced classification problem with an expected null accuracy of 50%. False positive rates (FPRs) and false negative rates (FNRs) are also provided for greater insight into model performance. Sub-analyses of accuracy in four different age groups (28 < 31, 31 < 34, 34 < 37 and 37 < 40 weeks) were performed in the Oxford Training Dataset (using the cross-validation results) and UCL Dataset only, as the subgroups in the Oxford Held-out Test Set were too small to generate reliable results (n = 4 in the youngest age group).

The permutation-based feature importance was estimated both in the out-of-bag samples in the final model derived from the Oxford Training Dataset (to assess feature importance in the training set) and on the Oxford Held-out Test Dataset (to assess features important for generalisation). The out-of-bag feature importance was calculated using a built-in functionality in the TreeBagger MATLAB function which calculates the decrease in out-of-bag classification accuracy if the variable is permuted across the samples that are out-of-bag for a given tree. For each feature, this value is standardised by averaging across all trees and divided by the standard deviation over all trees in the model. Feature importance in the Oxford Held-out Test Dataset was calculated in a similar way, by permuting the observations in the held-out set 50 times for each tree in the model separately for each feature. Mean accuracy loss across the 50 permutations was calculated for each feature and each tree and then averaged and standardised across all trees in the model. To further assess the power of the most important features, a model based on only the four most important features and PMA was trained across all available data, combining the Oxford Training Dataset, the Oxford Held-out Test Dataset and the UCL Dataset and accuracy was assessed by leave-one-infant out cross-validation.

#### Part 2: Development – developmental trajectories

To assess the developmental trajectories of the features included in the classification model, the data from the Oxford Training Dataset, Oxford Held-out Test Dataset and the UCL Dataset were combined, and all statistics were calculated across the three datasets. In the figures, the Oxford dataset (consisting of the Oxford Training Dataset and the Oxford Held-out Test Dataset) and the UCL Dataset are plotted in different colours in the same axes to show the similarity of the data across the two different research sites.

Developmental trajectories of the ERP, heart rate, limb withdrawal and ERSP features were qualitatively assessed by plotting age-weighted average responses in 4-week sliding windows. Within the window, each infant’s data were weighted so that infants with a PMA close to the centre PMA were upweighted and infants with a PMA further from the centre PMA were downweighed. A gaussian window was used to assign the weights between 0 and 1, so that infants who were one week from the centre PMA were assigned a weight of approximately 0.5 (see Supplementary Fig. S2). Developmental trajectories of brow bulge were expressed as the proportion of infants showing a brow bulge response (defined as a brow bulge duration above 0 seconds) in 3-week sliding windows.

The development of the subset of noxious-related features with PMA was also quantitatively assessed. The associations between PMA and heart rate, PC 1 noxious, PC 2 noxious, PC 3 noxious, Early Delta, Early Alpha, Late Delta, Late Alpha, Late Beta, ipsilateral reflex and contralateral reflex were quantified using linear regression models with p-values derived non-parametrically in the FSL PALM (permutation analysis of linear models) toolbox (Winkler et al. 2014) using 10,000 permutations. PALM was implemented in the MATLAB (Mathworks, version 2019b) environment. Each feature was tested separately, resulting in a total of 11 statistical tests and Holm-Bonferroni correction (Holm 1979) implemented in R (version 4.1.0, (R Core Team 2020)) was used to correct for multiple testing. The relationship between PMA and Brow bulge duration was not tested, as we previously reported these results for the Oxford dataset (Green et al. 2019); PC 1-3 non-noxious and PC 1-3 all data were not tested as these features are not extracted solely from the noxious data. Finally, the template was not tested because its waveform shows high similarity to PC3 noxious (see Fig. 4C).

Additional exploratory analyses were performed to investigate the effects that can more specifically be attributed to PNA or PMA adjusted for PNA by including PNA as a covariate in the linear models described above (see: Supplementary Table S1). As we were primarily interested in the effect of an infant’s overall age (gestational age + postnatal age) on the different response features, the adjustment for PNA was not used as the main analyses. These additional exploratory analyses are provided to give further insight into our primary PMA association results. Due to the exploratory nature of these analyses, the p-values are presented unadjusted for multiple testing.

Two-sided paired t-tests were used to compare the magnitude of PC 1-3 noxious between the noxious and the non-noxious stimuli with p-values derived non-parametrically in the FSL PALM (permutation analysis of linear models) toolbox (Winkler et al. 2014) using 10,000 sign-flips. Holm-Bonferroni correction was used to correct for multiple testing across these three tests.

To examine the clustering of features, each feature was first normalised to z-scores (subtraction of mean and division by standard deviation) within feature across all ages, then an age weighted average of the normalised feature was calculated at each PMA in 4-week sliding windows as described above. Linear models were used to obtain estimates for the regression coefficient between PMA and the z-scores and features were sorted by the magnitude of the regression coefficient. Clusters of features with similar trajectories were visually identified.

#### Software

Shaded plots were created using the shadedErrorbar function implemented in MATLAB (Campbell 2021).

## Results

### Neonates from 28 postmenstrual weeks have discriminable responses to noxious and non-noxious stimulation

Acute noxious stimulation in neonates from 28-40 weeks PMA evokes changes in brain activity, limb withdrawal activity, heart rate and facial expressions that can be discriminated from responses to non-noxious stimulation (Fig. 1). By averaging activity across this age range, visual inspection of the data confirms that noxious-evoked brain activity can be characterised by two distinct event-related potentials. The first N-P complex is present in response to both noxious and non-noxious stimulation, whereas the second negative-positive (N-P) complex is evoked solely by the noxious stimulation. Event-related spectral perturbation analysis demonstrates an increase in delta, alpha and beta power in the 2-second period post stimulation, which is greater in response to noxious stimulation compared with the non-noxious condition. In addition, an increase in heart rate and limb withdrawal activity are observed, which is greater following the noxious stimulation, and a greater proportion of neonates display facial grimacing (brow bulge) to the noxious stimulation (66 %) compared to the control condition (25 %).

**Figure 1.**
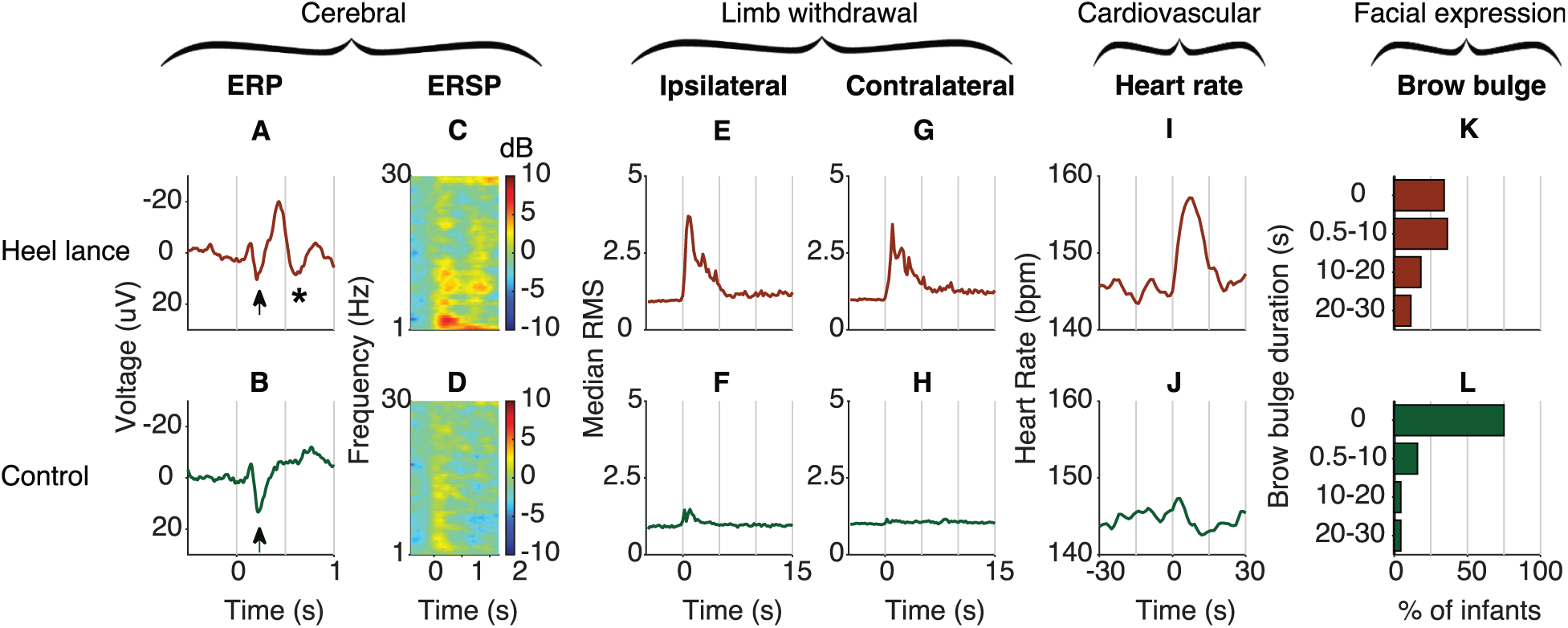
Average response to the noxious stimulus (heel lance, top row) and non-noxious stimulus (control, bottom row) in the Oxford Training Dataset (n = 47, infants aged 28-40 weeks PMA) for the cerebral, limb withdrawal, cardiovascular and facial expression modalities. The stimulus took place at time = 0 seconds. A-B) Mean ERP at the Cz electrode. An early N-P complex is visible in both the noxious and non-noxious responses (arrows), whereas the later N-P complex is only present in response to the noxious stimulus (asterisk). C-D) Mean ERSP plot. E-F) Median root-mean-square (RMS) of the ipsilateral limb withdrawal activity, plotted as fold-increase over a 1-second baseline. G-H) Median RMS of the contralateral limb withdrawal activity. I-J) Mean heart rate. K-L) Duration of brow bulge, shown as the percentage of infants displaying a brow bulge of a certain duration. Abbreviations: ERP = event-related potential; ERSP = event-related spectral perturbations.

A classification model, which was created based on the observed noxious-evoked cerebral, limb withdrawal, cardiovascular and facial expression responses (see Table 2 in Methods for the definition of each feature), had a discriminative accuracy of 0.79 (95% CI: 0.70-0.87) between the noxious and non-noxious conditions and an FPR of 0.28 and FNR of 0.15 in the Oxford Training Dataset (accuracy across all ages, estimated using leave-one-infant-out cross-validation; Fig. 2A-B). The model performed equally well in the independent Oxford Held-out Dataset, with an accuracy of 0.77 (95% CI: 0.65-0.89), FPR of 0.21 and FNR of 0.25, showing generalisability across datasets collected at the Oxford site.

**Figure 2.**
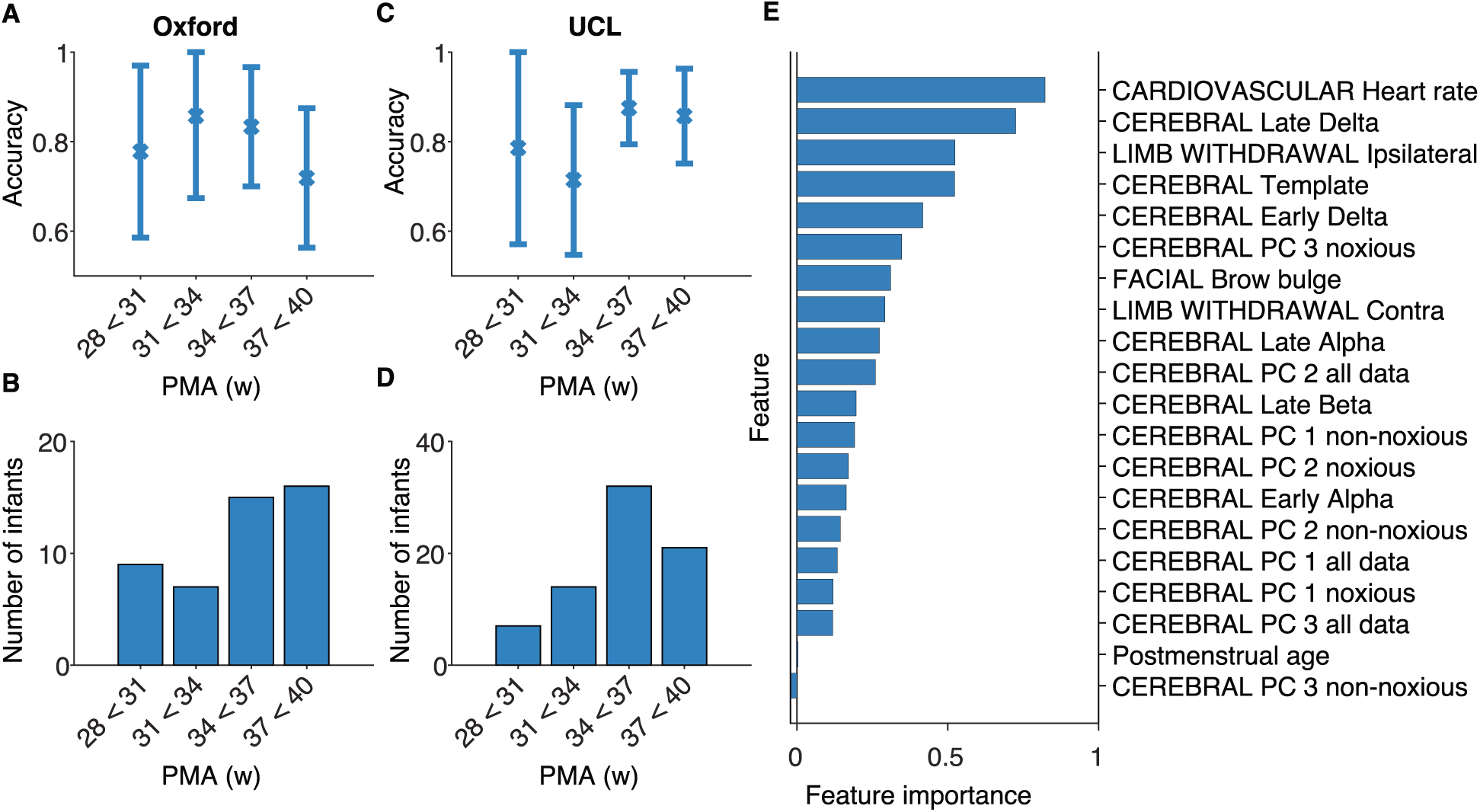
Overview of classification performance and feature importance estimates. A) Accuracy of the classification model across age in the Oxford Training Dataset (estimated using leave-one-subject-out cross-validation). Error bars represent 95% confidence interval. B) Number of infants in each age group in the Oxford Training Dataset. C) Accuracy of the classification model across age in the UCL Dataset. D) Number of infants in each age group in the UCL Dataset. E) Feature importance estimates obtained by permuting the observations in the Oxford Held-out Test Dataset. See Table 2 in the Methods for a description of each feature. Abbreviations: PC = principal component; PMA = postmenstrual age.

Importantly, these results were confirmed in another independent dataset collected by a different research group at a different research site (UCL Dataset (Jones et al. 2018b)), demonstrating the generalisability of these findings (Fig. 2C-D). The accuracy of the classifier in the UCL Dataset was 0.83 (95% CI: 0.77 – 0.89), with an FPR of 0.11 and an FNR of 0.23.

Differences in accuracy across age were investigated in the Oxford Training Dataset and the UCL Datasets, where sufficiently large subgroups could be generated based on PMA (Fig. 2A,C). The accuracy was balanced across age groups. Notably, in the 28-31 week group the accuracy was 0.78 (95% CI 0.59 – 0.97) in the Oxford Training Data and 0.79 (95% CI 0.57 – 1.00) in the UCL Dataset, confirming that neonatal responses to noxious and non-noxious stimuli can be discriminated in very preterm infants.

The importance of each feature to the model was evaluated, which showed that the heart rate, Late Delta activity, ipsilateral limb withdrawal and the template of noxious-evoked brain activity gave the greatest contribution to the classification accuracy in both the Oxford Training Dataset and the Oxford Held-out Test Dataset (Fig. 2E, Fig. S3). These features span cardiovascular, behavioural and cerebral modalities, demonstrating that classification benefits from data recorded across multiple domains. A new classifier trained on only these four features and PMA across all available data (the Oxford Training Dataset, Oxford Held-out Test Dataset and the UCL Dataset) performed equally well with an overall discriminative accuracy of 0.79, estimated using leave-one-out cross-validation (95% CI 0.74-0.84, varying from 0.71 in the 31-34 weeks PMA group to 0.81 in the 37-40 weeks PMA group).

### Noxious-evoked cerebral, limb withdrawal, cardiovascular and facial expression responses are age dependent

Distinct morphological changes in the noxious-evoked activity can be observed between 28-40 weeks PMA, which are highly consistent across the independently collected Oxford and UCL datasets (Fig. 3). To quantititatively investigate age-related changes in noxious-evoked responses, the Oxford Training Dataset, Oxford held-out Test Dataset and UCL Dataset were combined. Noxious-evoked increases in heart rate are small in the youngest infants and significantly increase with PMA (Fig. 3B, n = 131, beta = 1.66, t = 7.07, Holm-corrected p = 0.0011). In contrast, the magnitude of the withdrawal reflex in either the ipsilateral or contralateral limb in the first second post-stimulus does not significantly change with age (Fig. 3C; ipsilateral limb: n = 69, beta = −0.063, t = −0.34, Holm-corrected p = 1.0; Fig. 3D; contralateral limb; n = 68, beta = 0.23, t = 1.03, Holm-corrected p = 1.0), although greater magnitude responses are visually apparent in the younger infants. The proportion of infants who showed facial grimacing to the noxious stimulus increases from 43 % in the youngest age group to 60 % in the oldest age group in the Oxford Dataset as we previously reported in (Green et al. 2019) and from 40 % to 64 % in the UCL Dataset (Fig. 3E).

**Figure 3.**
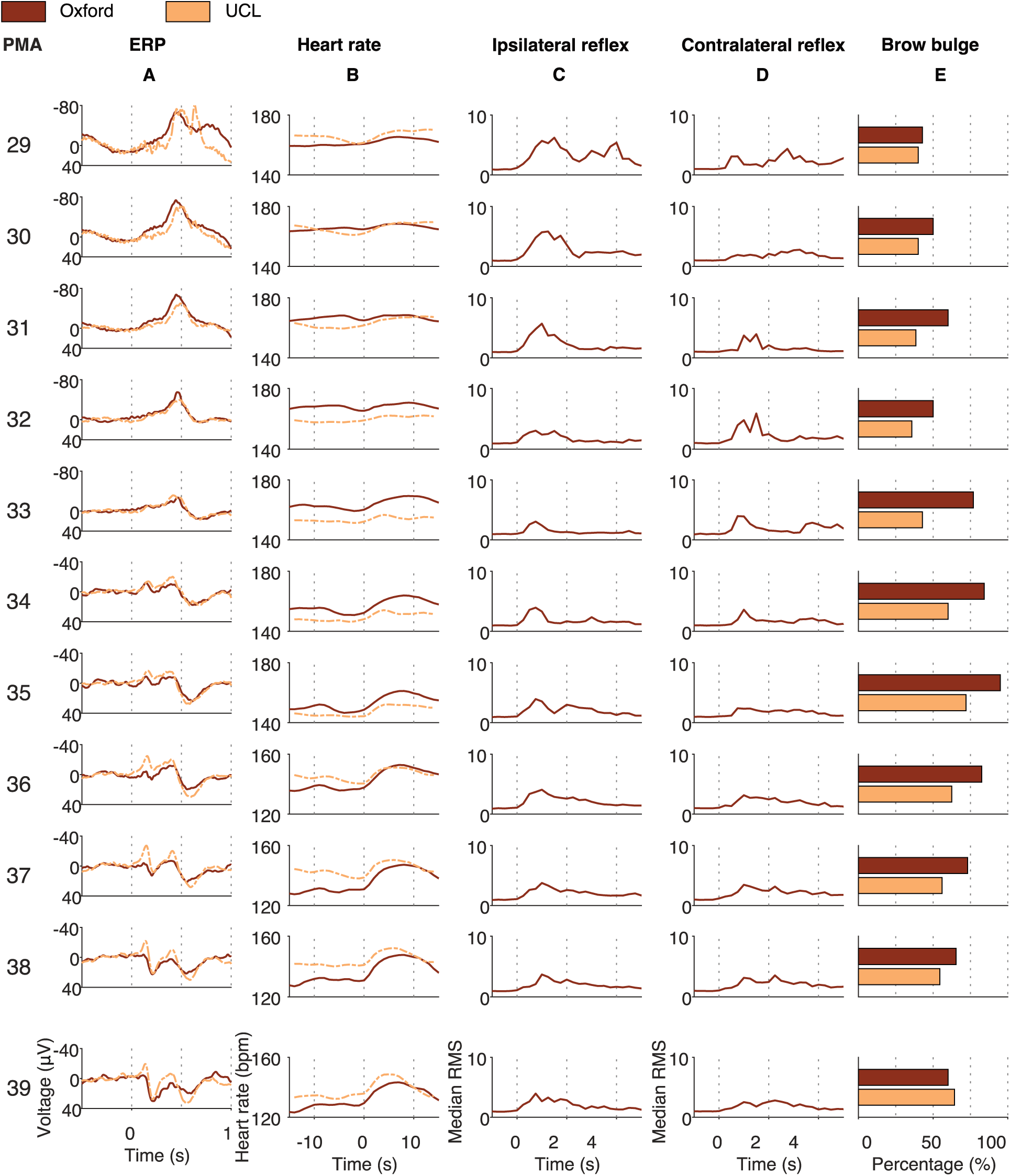
Noxious-evoked ERP, heart rate, ipsilateral reflex, contralateral reflex and brow bulge responses in neonates from 28-40 postmenstrual weeks, split by postmenstrual week. For the continuous variables, each trace is an age-weighted average in 4-week sliding windows around the centre PMA. For the brow bulge responses, bars demonstrate the percentage of infants that displayed a brow bulge response in a group of infants with a PMA that falls within 1.5 weeks relative to the centre PMA. Oxford data contains the Oxford Training Dataset and the Oxford Held-out Test Dataset. EMG is not available in the UCL Dataset. Column A) ERP morphology changes from a large negative peak in 30-33 week PMA to a positive waveform. Column B) Heart rate rise increases with PMA. Columns C-D) Noxious-evoked reflex activity in the first second post-stimulus does not significantly change across development. Ipsilateral reflex responses visually decrease in magnitude when examining the entire reflex duration. Column E) The proportion of infants displaying a brow bulge response increases with PMA. Abbreviations: ERP = event-related potential; PMA = postmenstrual age.

**Figure 4.**
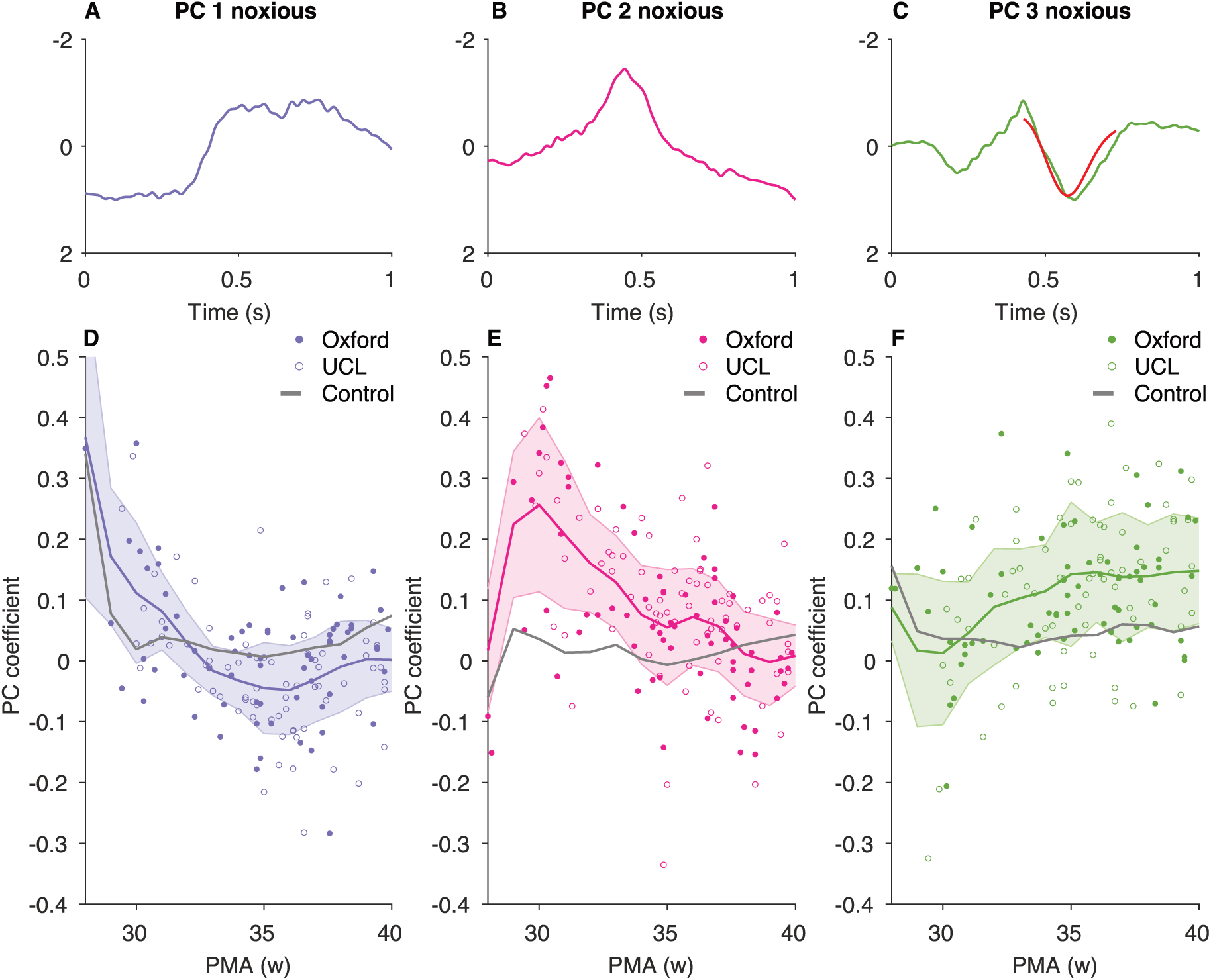
The three PCS derived in the noxious data in the Oxford Training Dataset and their associated age-related trajectories in the Oxford and UCL datasets. A-C) The three PCs derived from the noxious data in the Oxford Training Dataset. In (C), the previously published template of noxious-evoked brain activity (Hartley et al. 2017) is scaled and overlaid in red to show the similarity of the waveforms. D-F) Corresponding age-related trajectories of the PC coefficients. PC coefficients were obtained by projecting the PCs to the Oxford Training Dataset, Oxford Held-out Test Dataset and UCL Dataset. Dots represent individual infants (noxious responses only). Developmental trajectories (colour and grey lines: noxious and non-noxious trajectory, respectively) are estimated for visualisation only by calculating the age-weighted average and standard deviation of the PC coefficients at each postmenstrual week (see Methods). Oxford data contains the Oxford training set and the Oxford Held-out Test Dataset. Abbreviations: PC = principal component; PMA = postmenstrual age.

In the youngest infants, the time-locked EEG activity to both noxious and non-noxious stimulation consists of a high-amplitude slow wave superimposed with high-frequency activity, resembling previously-reported delta brush activity (Fabrizi et al. 2011). From 30 weeks PMA a prominent ERP is evoked by the noxious stimulation, which has not been previously reported, and consists of a single negative peak at approximately 400-500 ms that decreases in magnitude with increasing PMA (Fig. 3A) and is not present in response to the non-noxious procedure (Supplementary Fig. S4). From 33 weeks onwards, a positive peak at approximately 500-600 ms is evoked that resembles previously well-characterised noxious-evoked activity (Slater et al. 2010; Fabrizi et al. 2011).

### The morphology of noxious-evoked brain activity changes throughout early development

To investigate further the morphology of the developing noxious-evoked brain activity, we examined the presence of the main waveforms that were identified in the noxious responses in the Oxford Training Dataset. The first three waveforms (PC 1, PC 2 and PC 3, Fig. 4) account for over 80 % of the variance in the noxious-evoked activity in the Oxford Training Dataset and represent the activity that can be visually observed across distinct developmental stages (Fig. 3). PC 1 resembles the slow wave component of previously reported delta brush activity (Fabrizi et al. 2011); PC 2 is a negative deflection with peak latency at approximately 445 ms; and PC 3 is composed of an early positive and negative deflection (latency: 214 ms and 429 ms respectively) followed by a second positive deflection peaking at approximately 595 ms and resembling the previously published template of noxious-evoked brain activity in term infants (Fig. 4C (Hartley et al. 2017)). When the PCs are projected onto the Oxford and UCL Datasets they discriminate between the noxious and non-noxious conditions; PC 2 and PC 3 are significantly greater in response to the noxious stimulation compared with non-noxious stimulation (paired t-test, PC 2; n = 145, t = −5.32, Holm-corrected p = 0.0003, PC 3; n = 145, t = −5.15, Holm-corrected p = 0.0003), while PC 1 is significantly greater following non-noxious stimulation (paired t-test, n = 145, t = 2.73, Holm-corrected p = 0.0083).

The PC coefficients follow distinct age-related trajectories (Fig. 4). PC 1 is dominant in the neonates aged 28-29 weeks PMA and significantly decreases in magnitude with increasing PMA (n = 145, beta = −0.016, t = −5.16, Holm-corrected p = 0.0011). This coincides with the appearance of PC 2, which peaks at approximately 30 weeks PMA and steadily decreases until term age (n = 145, beta = −0.022, t = −6.74, Holm-corrected p = 0.0011). Meanwhile, PC 3 increases in magnitude with age (n = 145, beta = 0.012, t = 3.75, Holm-corrected p = 0.0024) and reaches a plateau at term age. The results are similar when correcting for PNA (see Supplementary Table S1).

The ERSP features show weaker associations with age (Fig. 5). Of the 5 ERSP features that were visually identified in response to the noxious stimulation in in the Oxford Training Dataset (Fig. M1, Table 3), only Early Alpha significantly changes with PMA (n = 67, beta = −0.55, t = −3.14, Holm-corrected p = 0.019). The other features are relatively constant in magnitude.

**Figure 5.**
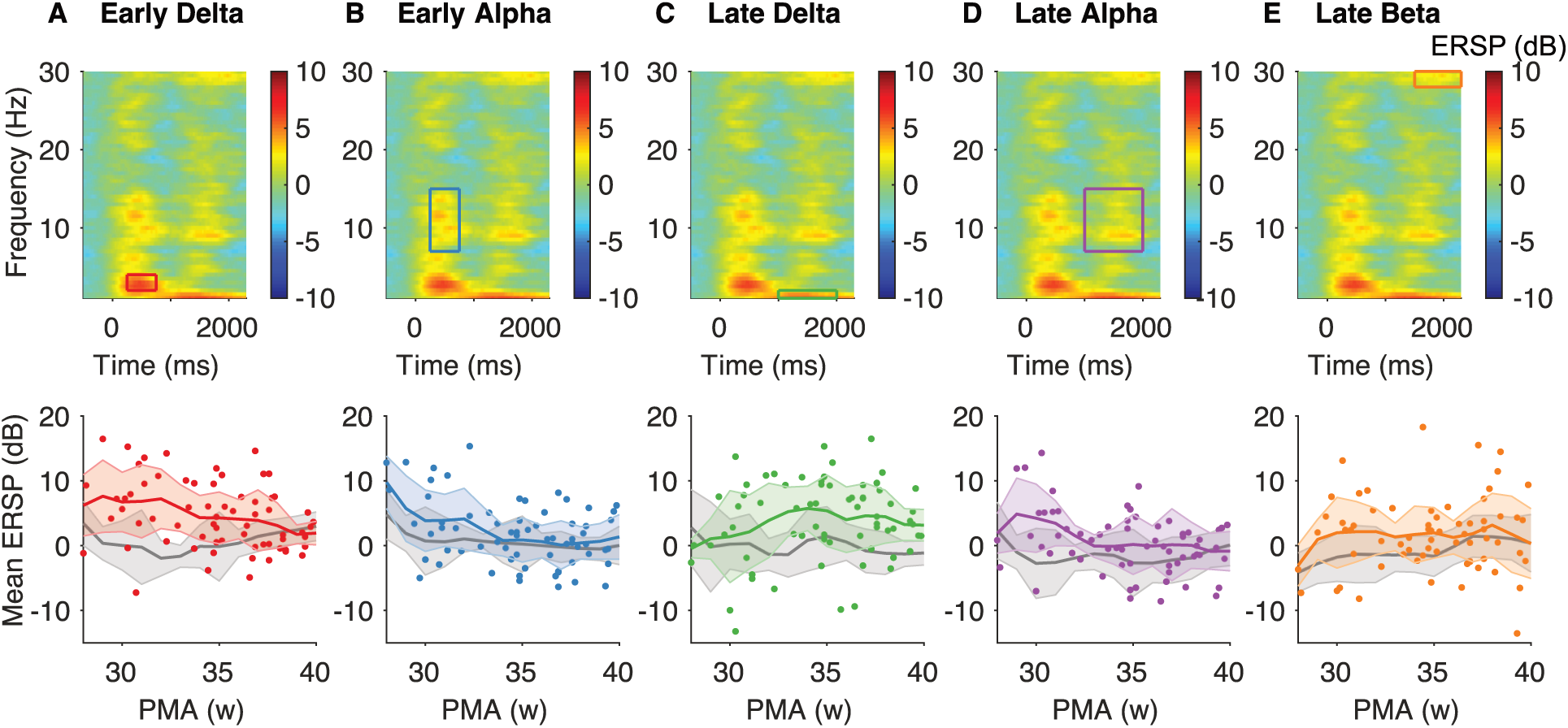
Age-related changes in the five ERSP features. Top row: Average ERSP response to the noxious stimulus in the Oxford data with time-frequency windows of interest: Early Delta (A), Early Alpha (B), Late Delta (C), Late Alpha (D) and Late Beta (E). The stimulus occurred at time = 0. Bottom row: Corresponding developmental trajectories of the ERSP features. Dots represent individual infants (noxious responses only). Colour and grey lines represent the mean noxious and non-noxious trajectory, respectively, and are estimated for visualisation only by calculating the age-weighted average and standard deviation of the ERSP feature magnitude at each postmenstrual week (see Methods). Oxford data contains the Oxford training set and the Oxford Held-out Test Dataset. Abbreviations: ERSP = event-related spectral perturbations; PMA = postmenstrual age.

### The pattern of noxious-evoked multimodal activity changes throughout early development

The maturation of the responses across the different modalities follows differing developmental trajectories (Fig. 6). Two clusters of responses are visually apparent. PC 2 noxious, PC 1 noxious, Early Alpha, Early Delta and Late Alpha are greater in infants below 33 weeks and decrease in magnitude with increasing PMA. In contrast, heart rate increase, PC 3 noxious, brow bulge duration, the magnitude of the pre-defined template of noxious-evoked activity and Late Delta, are greater in infants above 36 weeks and smaller and infants below 33 weeks.

**Figure 6.**
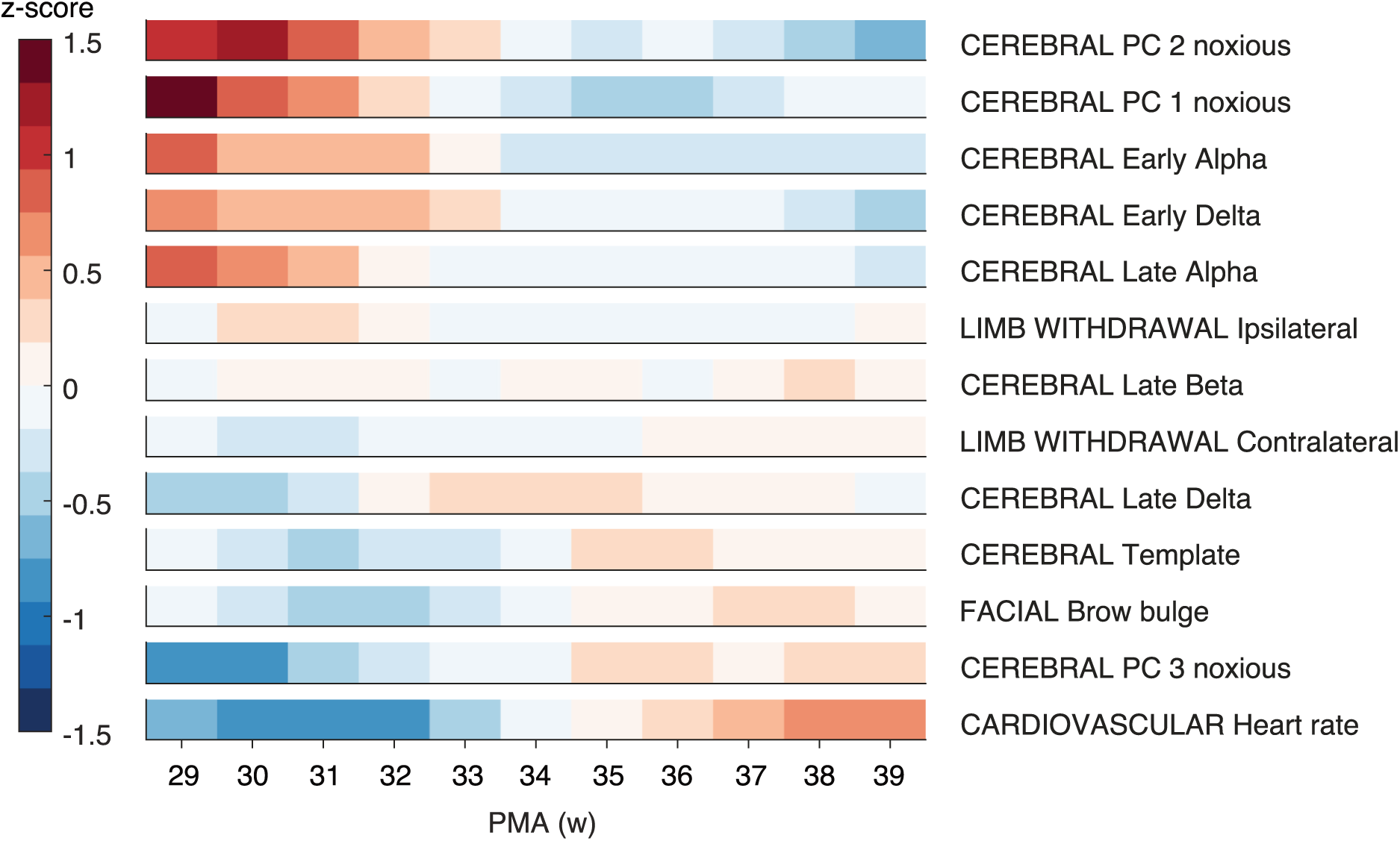
Normalised magnitude of noxious-related responses at increasing postmenstrual weeks, separated by feature. Magnitudes were first normalised within modality across all infants, then age-weighted averages were calculated in a 4-week sliding window where infants close to the centre PMA were upweighted (see Methods). Abbreviations: PMA = postmenstrual age.

## Discussion

Characterising noxious-evoked neonatal responses to potentially painful medical procedures is crucial to better measure and treat pain in hospitalised neonates. The youngest premature neonates, born from approximately 22 weeks’ gestation, experience the highest number of clinically-necessary interventions per day (Carbajal et al. 2008), which occur during a developmental period where they are vulnerable to long-term detrimental consequences (Doesburg et al. 2013; Ranger et al. 2015; Duerden et al. 2018). While there are many lines of evidence to suggest that term-aged infants display discriminable patterns of behavioural, autonomic and cerebral activity in response to noxious events, in contrast, in premature infants a lack of discrimination between observed responses to noxious and non-noxious stimulation is often reported. Here we demonstrate that neonates from 28 weeks’ PMA display developmentally distinct activity patterns that discriminate between noxious and non-noxious stimuli. Using a classification model, we show that multimodal activity, including evoked brain activity, behavioural responses and autonomic responses, can discriminate noxious from non-noxious stimuli with an accuracy of 77 to 83%. Importantly, the accuracy was 78-79% in the youngest age group (28-31 weeks PMA) demonstrating that even in infants of this age it is possible to discriminate the responses to the two stimuli.

In this study we demonstrate that evoked brain activity patterns recorded using EEG change morphology across the developmental period from 28-40 weeks PMA. Notably, we report the emergence of a transitory noxious-evoked waveform in infants aged approximately 30-33 weeks PMA that has not previously been described. Identification of this transitory activity, consisting of a negative deflection at approximately 400-500 ms following the noxious stimulus, has the potential to facilitate the use of noxious-evoked activity as a surrogate pain outcome measure in research investigations and clinical trials in neonates aged less than 34 weeks PMA, analogous to the use of a standardised template of noxious-evoked brain activity in term infants (Hartley et al. 2017, 2018). Additionally, we confirm previously reported observations that noxious stimulation evokes immature delta brushes in very premature infants (Fabrizi et al. 2011) and that from approximately 34 weeks PMA a previously well-characterised pattern of noxious-evoked activity emerges consisting of a positive deflection at approximately 500 ms following the noxious stimulus (Slater et al. 2010; Hartley et al. 2017). The observed changes in the noxious-evoked brain activity pattern follow a continuous developmental transition, which could potentially underpin differences in emotional and sensory perceptions evoked by the nociceptive input at different developmental ages. In contrast to the other modalities, where discrimination between the noxious- and the non-noxious procedure is based on magnitude, ERP responses are qualitatively different between noxious and non-noxious stimulation. Whilst in the absence of verbal report we do not know how infants perceive these stimuli, importantly, identification of morphologically distinct noxious-evoked brain activity from 30 weeks PMA highlights the possibility of infants differentially processing noxious and non-noxious stimuli at this age, and emphasises the need for appropriate pain management for all infants.

The developmental period studied from 28-40 weeks PMA coincides with major changes in the structural maturation of nociceptive pathways. Between 26 and 28 weeks’ gestation, thalamo-cortical fibres innervate somatosensory areas in the cortical plate through the subplate, a transient structure underlying the cortical plate (Kostovic and Judaš 2010; Krsnik et al. 2017). Between 31 and 34 weeks’ gestation the cortical plate differentiates into its adult-like configuration with six layers (Kostovic and Judaš 2010). From approximately 35 weeks’ gestation the subplate begins to disappear, giving way to mature thalamo-cortical connections, and inter-hemispheric connections start to form (Kostovic and Judaš 2010). Considering these changes, the emergent transitory brain activity may represent early thalamo-cortical signalling, whereas the more mature later positive waveform may represent further processing involving inter- and intra-hemispheric connections between the cortex and other brain areas. This would be in line with findings in adults where the individual components of noxious-evoked ERPs covary with activity in distinct brain regions (Mobascher et al. 2009) and each mediate different aspects (perceptual, motor or autonomic) of the pain-related response (Tiemann et al. 2018). An important next step is to investigate the development of the spatial organisation of the noxious-related brain activity in preterm infants using techniques such as fMRI, which have successfully been used in term infants (Goksan et al. 2015; Williams et al. 2015; Duff et al. 2020). Importantly, the emergence of remarkably similar patterns of noxious-evoked activity have also been observed in rat pups after sensory fibre stimulation (Chang et al. 2020), facilitating the translation between laboratory and clinical investigations.

Besides temporal EEG changes, we identified changes in EEG event-related spectral perturbations in response to noxious stimulation. In line with previous findings (Fabrizi et al. 2016), we observed an increase in power mostly in the delta, alpha and high-beta ranges. Late Delta (1-2 Hz, 1-2 seconds post-stimulus) was an important discriminative feature in our classification model, while the higher frequency ERSP features – High Beta (28-30 Hz) and Early and Late Alpha (7-15 Hz) – had lower feature importance. This is in apparent contrast with some investigations in adults, where painful stimulation is reported to be mainly associated with decreases in alpha and beta activity and increases in gamma activity (Tu et al. 2016; Misra et al. 2017; Ploner et al. 2017). Pain-related responses in preterm infants may be of lower frequency than in adults, considering that the preterm EEG generally has a lower frequency content than the adult EEG (Niemarkt et al. 2011) and adult EEG frequency band cut-offs cannot be directly translated to infants (Saby and Marshall 2012). It would be of interest to further investigate the development of noxious-evoked ERSPs in infants at higher postnatal ages. Future work should also include multiple electrodes to investigate the spatial distribution of these responses.

The maturational development of the autonomic and behavioural responses was also apparent in this study. The evoked change in heart rate and brow bulge increased in magnitude between 28 and 40 weeks PMA. Both responses matured concomitantly with the appearance of the more mature noxious-evoked brain activity that has been well-characterised in term-aged infants. This is in line with previous work which suggested that discrimination in facial expression responses between noxious and non-noxious stimuli is related to the maturity of the brain activity response (Green et al. 2019). Interestingly, the youngest infants here only display a small increase in heart rate to the noxious stimulus compared to term infants, which is apparent in both independently collected datasets. While a limitation of the study is the relatively small numbers of infants at the youngest ages (13 infants in the Oxford dataset and 7 infants in the UCL Dataset were aged less than 31 weeks PMA), this dependence of the magnitude of the heart rate response on PMA highlights the need for developmentally adjusted clinical pain assessment.

The magnitude of the noxious-evoked reflex activity did not seem to attenuate with age. However, this apparent discordance with previous work (Cornelissen et al. 2013; Hartley et al. 2016) may be explained by the relatively short time window (1 second) of response that was used for the analysis - it seems that while the initial magnitude of the rise of the reflex is relatively constant with age, the duration of the responses appear visually longer in the younger neonates, consistent with previous reports (Hartley et al. 2016). Future work to refine the model presented here should investigate whether different characteristics of the limb withdrawal could be used at different ages to further improve discrimination accuracy. Importantly, the classification and feature importance results derived from the training data generalised to a statistically independent test set. The most important features spanned cardiovascular, cerebral and limb withdrawal modalities in both the training and testing data, confirming the importance of multimodal assessments. Moreover, the classification results and developmental changes in noxious-evoked activity were reproduceable in a fully independent dataset collected at a different centre by a different team of researchers (Jones et al. 2018b).

This demonstrates the consistency of these techniques and the utility of brain-derived approaches to better understand how painful procedures impact the activity of the central nervous system in developing infants. Although implementing these techniques in new centres can be challenging (Baxter et al. 2021), the development of a clinically useable tool with simplified EEG acquisition and integration of standard analysis techniques will facilitate the use of these methods for the measurement of noxious-evoked brain activity in research investigations and clinical trials.

The goal of the current study was not to develop a clinical tool with the highest achievable classification accuracy, but to investigate discrimination across age. While we achieved reasonably high classification accuracy, further investigations into alternative algorithms, feature selection and hyperparameter optimisation could facilitate the creation of a clinically useable tool to measure noxious-evoked cerebral, behavioural and autonomic activity in infants from 28 weeks PMA that is developmentally sensitive. In addition, the developmental trajectories described here could be further modelled to specifically investigate the transitions in multimodal patterns of activity and to determine the normal boundaries for interpreting noxious-related responses at different ages. As this study included only one type of noxious stimulation (a heel lance), future studies should address the similarity of responses evoked by other clinical procedures, such as immunisations or cannulations. In this work, the noxious and non-noxious stimulus were closely matched in terms of tactile, vibratory and auditory aspects. To further investigate the specificity of the noxious-evoked responses, it would also be of interest to investigate the discrimination between noxious procedures and other non-noxious modalities (e.g., visual) that are matched in saliency (Mouraux and Iannetti 2018).

In summary, we demonstrate that evoked responses to noxious and non-noxious stimuli can be discriminated in infants from 28 weeks PMA. We characterise the developmental changes in brain activity, behavioural and autonomic responses between 28-40 weeks PMA, including the transient emergence of a previously uncharacterised pattern of noxious-evoked brain activity observed between approximately 30-33 weeks PMA. Whilst developmental changes with distinct features occur across this age range, responses can be discriminated at every age. Better understanding of the response to noxious stimuli with age will lead to the construction of developmentally sensitive clinical pain measurement tools and more accurate assessment of the impact of analgesic interventions.

## Data Availability

Data from the Oxford Dataset are available from the corresponding author upon reasonable request. Due to ethical restrictions, it is appropriate to monitor access and usage of the data as it includes highly sensitive information. Data sharing requests should be directed to.
The UCL dataset is a published dataset collected by an independent research team which is available through the UK Data Service. Data reference: Jones L, Laudiano-Dray M, Whitehead K, Verriotis M, Meek J, Fitzgerald M, Fabrizi L. 2018. EEG, behavioural, and physiological responses to a painful procedure in human neonates with relevant medical history [data collection]. UK Data Service SN: 853311, http://doi.org/105255/UKDA-SN-853204. Dataset description and methods: Jones et al. Sci Data. 2018;5:180248.

## Acknowledgements

We would like to thank Laura Jones and Lorenzo Fabrizi for sharing the UCL Dataset with us through the UK Data Service. We would like to thank all other members of the Paediatric Neuroimaging group for help with data acquisition. We are grateful to all the families who participated in the studies.

