## Supplementary materials for "Premature infants display discriminable behavioural, physiological and brain responses to noxious and non-noxious stimuli"

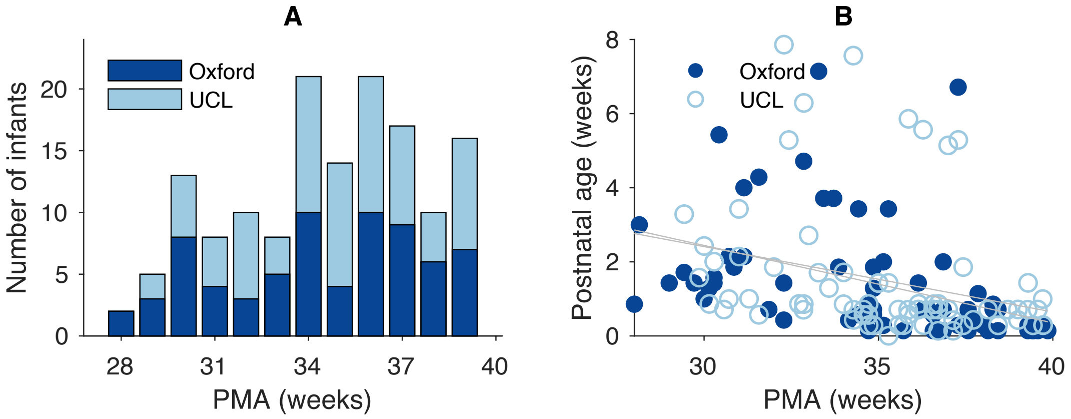


**Figure S1. Age distribution of the Oxford and UCL Datasets.** A) Distribution of postmenstrual age (PMA) in the Oxford Dataset (combined Training and Held-out Test Dataset) and UCL Dataset. B) Relationship between PMA and postnatal age. Grey lines are the lines of best fit in the two datasets.

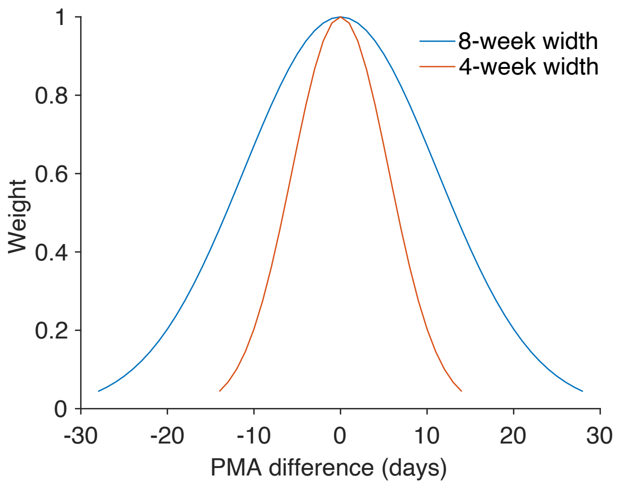


**Figure S2.** Gaussian windows used to assign weights for the calculation of age-weighted averages before the principal component analysis (8-week width) and the age trajectories presented in Figures 4-7 (4-week width).

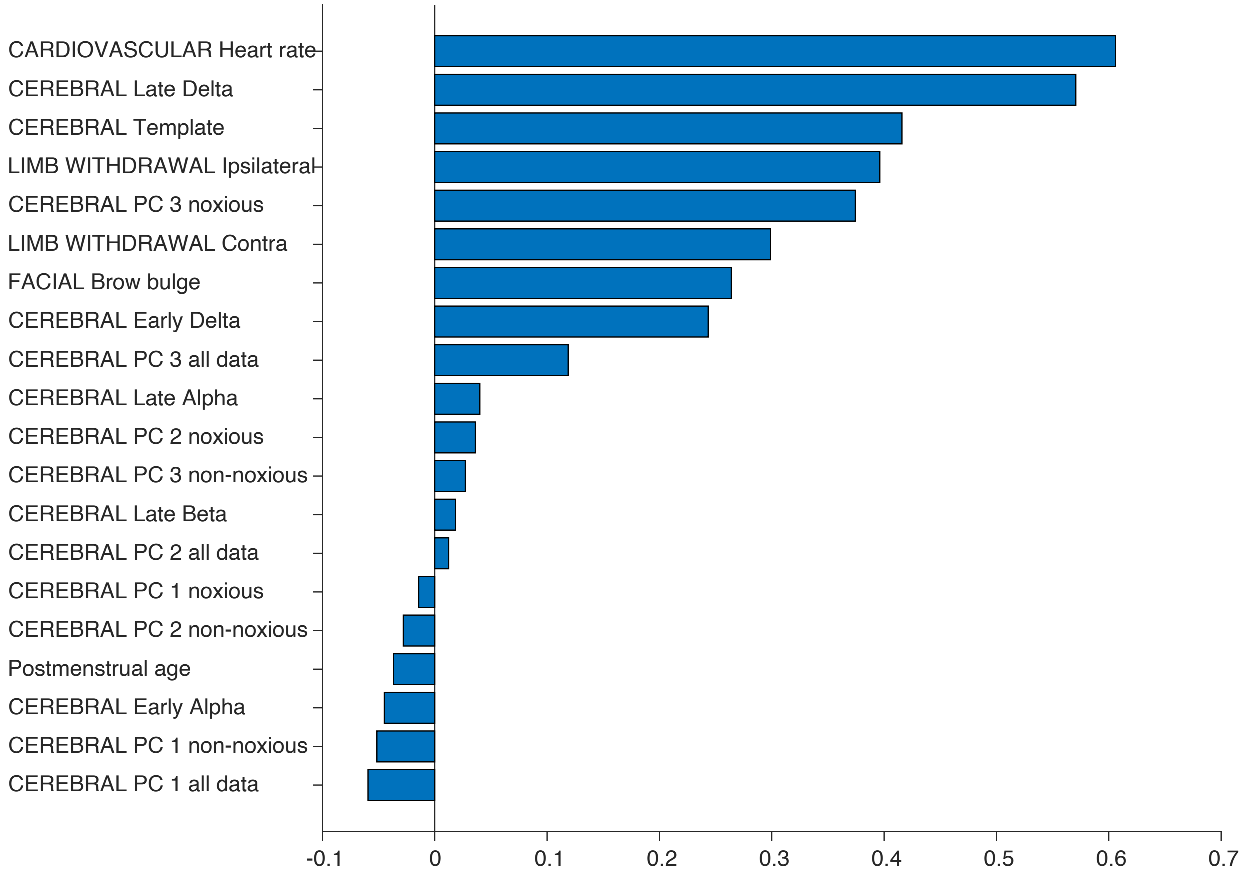


**Figure S3.** Permutation feature importance estimated in the out-of-bag samples in the training data. Abbreviations: PC = principal component.

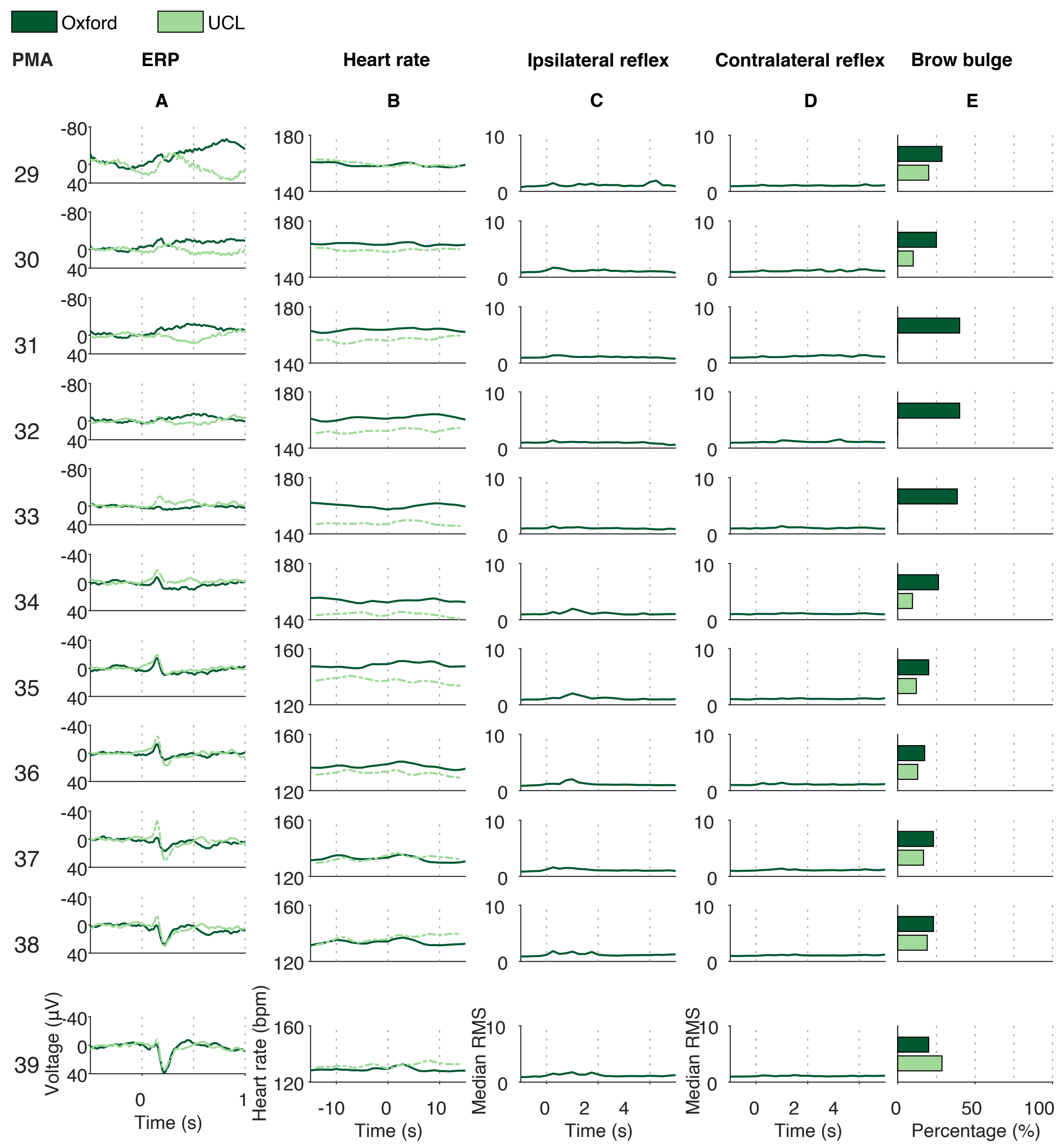


**Figure S4. ERP, heart rate, ipsilateral reflex, contralateral reflex and brow bulge responses to the non-noxious control procedure in neonates from 28-40 postmenstrual weeks, split by postmenstrual week.** For the continuous variables, each trace is an age-weighted average in 4-week sliding windows around the centre PMA. For the brow bulge responses, bars demonstrate the percentage of infants that displayed a brow bulge response in a group of infants with a

PMA that falls within 1.5 weeks relative to the centre PMA. Oxford data contains the Oxford training set and the Oxford Held-out Test Dataset. EMG is not available in the UCL Dataset. Column A) ERP. Column B) Heart rate. Columns C-D) Ipsilateral and contralateral reflex responses. Column E) Proportion of infants displaying a brow bulge response. Abbreviations: ERP = event-related potential; PMA = postmenstrual age.

**Table S1.** Regression coefficients and statistics for associations between (left) PMA and noxious-response metrics corrected for PNA, and (right) PNA and noxious-response metrics corrected for PMA. Noxious-response metrics are sorted by p-value for the PMA covariate. Results are consistent with the analyses in the main text. P-values are uncorrected for multiple testing due to the exploratory nature of the analyses. Abbreviations: PC = principal component.

| | PMA | | | PNA | | |
|---|---|---|---|---|---|---|
| | **Beta** | **t-stat** | **p** | **Beta** | **t-stat** | **p** |
| PC 1 | -0.017 | -5.25 | 0.00010 | -0.007 | -1.18 | 0.24 |
| PC 2 | -0.021 | -5.99 | 0.00010 | 0.006 | 0.96 | 0.33 |
| Heart rate | 1.60 | 6.40 | 0.00010 | -0.31 | -0.72 | 0.47 |
| PC 3 | 0.010 | 2.90 | 0.0041 | -0.012 | -1.87 | 0.066 |
| Early Alpha | -0.47 | -2.46 | 0.018 | 0.38 | 0.96 | 0.35 |
| Late Delta | 0.49 | 1.98 | 0.056 | 1.14 | 2.25 | 0.028 |
| Early Delta | -0.38 | -1.86 | 0.071 | 0.63 | 1.51 | 0.14 |
| Late Beta | 0.35 | 1.54 | 0.14 | 1.23 | 2.62 | 0.010 |
| Late Alpha | -0.22 | -1.05 | 0.31 | 1.08 | 2.60 | 0.014 |
| Ipsilateral reflex | -0.21 | -0.98 | 0.33 | -0.63 | -1.34 | 0.18 |
| Contralateral reflex | 0.20 | 0.77 | 0.44 | -0.19 | -0.36 | 0.72 |
